# Protective effects of sacubitril/valsartan in anthracyclines-induced cardiotoxicity: A Systematic Review and Meta-Analysis

**DOI:** 10.1101/2023.08.14.23294099

**Authors:** Felipe Israel López Trejo, Mónica Daniela Sánchez Barrera, Elias Noel Andrade Cuellar, Julieta Danira Morales Portano, Laura Aline Martínez Martínez

## Abstract

**Background:** One of the adverse effects of anthracycline use is cardiotoxicity manifested as heart failure. The benefit of sacubitril/valsartan in patients with heart failure of ischemic origin has been demonstrated, but not due to toxicity. Our objective was to determine if there are protective effects of the use of sacubitril/valsartan in cardiotoxicity induced by anthracyclines.

**Methods:** A bibliographic search was carried out in PubMed, Cochrane Library, LILACS, Scopus, EBSCO and Google Scholar. A systematic review and meta-analysis was performed to assess mean differences (MD) with 95% confidence intervals following the PRISMA Statement.

**Results:** 558 published articles were found, of which three were submitted to analysis, with a total sample of 69 patients. With the use of sacubitril/valsartan there was an increase in the percentage of left ventricular ejection fraction (LVEF) [MD 6.78% (p < 0.001)], an increase in the percentage of global longitudinal strain of the left ventricle (LV-GLS) [MD 3.37 % (p < 0.001)], and decreased serum levels of NT-proBNP [MD −1003.64 pg/mL (p < 0.001)]; in addition, there was an increase in the distance traveled with the use of sacubitril/valsartan [MD 95.23 meters (p < 0.001)] compared to their baseline values, respectively.

**Conclusions:** This meta-analysis suggests that there is a benefit from the use of sacubitril/valsartan in patients with heart failure related to the use of anthracyclines on ventricular remodeling and functional capacity. This is the first meta-analysis to our knowledge evaluating this effect. Randomized double-blind placebo-controlled clinical trials are required to confirm these findings.

## INTRODUCTION

Chemotherapies unwanted side effects, among which cardiotoxicity stands out, are causing patients die from cardiotoxicity and not from cancer-related causes^1^, being the cardiovascular disease currently the second leading cause of long-term morbidity and mortality among cancer survivors^2^. Chemotherapy-related cardiotoxicity is clinically manifested as heart failure^3^, being the anthracyclines the main group involved^4^. It is worrisome that only 11 % of patients who develop this complication recover myocardial function, leaving the remaining 89% with sequelae^5^. Left ventricular ejection fraction (LVEF) is the most commonly accepted parameter for evaluating cardiac function, as it independently predicts short- and long-term mortality from cardiovascular events, including anthracycline-induced cardiomyopathy^8^. Serum biomarkers have also shown utility as diagnostic and prognostic methods in this disease, mainly NT-proBNP^7^ and troponin I^6^. In the treatment of heart failure, the main therapeutic targets are blockade in the renin-angiotensin-aldosterone system (RAAS) and neprilysin inhibition (ARNI). The drug currently available that allows combined inhibition of the renin-angiotensin system and neprilysin without adverse events is sacubitril/valsartan^9^. The use of ARNI has shown superiority over other combination therapies^10^ in reducing mortality rates. The current guidelines of the American Heart Association (AHA)^11^ and the ESC^12^ recommend the use of four drugs that have been shown to reduce mortality and hospitalization rates as standard treatment for heart failure with reduced ejection fraction, including angiotensin-neprilysin receptor inhibitors (ARNI); however, the clinical trials that support the benefits of their use do not include cancer patients, so the use of these drugs is controversial in the context of chemotherapy-related cardiac dysfunction, and prevention and treatment strategies are needed to improve the clinical outcomes in these patients. For these reasons, we decided to carry out a systematic review and meta-analysis of the beneficial cardioprotective effects of the use of sacubitril/valsartan in the treatment of patients with anthracycline-induced cardiotoxicity, to provide more scientific information that supports or not the performance of randomized controlled trials, since there are no complete published studies to date, and there is no meta-analysis to our knowledge that evaluates this.

## METHODS

The primary objective of this study was to determine whether there are protective effects of the use of sacubitril/valsartan in the treatment of adult patients with anthracycline-induced cardiotoxicity. Secondary objectives were to determine the effects of sacubitril/valsartan on echocardiographic variables of myocardial function, on serum biomarker levels, and on drug safety variables (serum creatinine and potassium levels).

A systematic search was performed in six databases until July 2022: PubMed, Cochrane Library, LILACS, Scopus, EBSCO and Google Scholar, with four search algorithms by two reviewers: [cardiotoxicity AND anthracyclines AND sacubitril and valsartan sodium hydrate drug combination], [cardiotoxicity AND anthracyclines AND sacubitril], [cancer therapy-related cardiac dysfunction AND sacubitril and valsartan sodium hydrate drug combination] and [cancer therapy-related cardiac dysfunction AND sacubitril].

The inclusion criteria were articles that include individuals older than 18 years, with cancer, who had received treatment with an anthracycline at any cumulative dose, who presented chemotherapy-related myocardial dysfunction as a manifestation of cardiotoxicity, and who had received treatment with sacubitril/valsartan at any dose. The exclusion criteria were individuals who had presented heart failure prior to their inclusion in the corresponding prospective study and individuals with follow-up of less than three months.

A systematic review and meta-analysis was performed based on the Cochrane methodology for non-randomized studies. Data are expressed as means and standard deviation or interquartile ranges for quantitative variables according to their distribution. Qualitative variables are expressed in percentages or proportions. Inverse variance was used to measure the effect. Confidence intervals were set at 95 %. Results are expressed as mean difference. The fixed effects analysis model was used. Only in cases of significant heterogeneity was the random effects model used. Tests for heterogeneity between studies were performed using the Cochran Q test^13^ and the I^2^ test^14^. A p value < 0.05 or an I^2^ greater than 70 % were considered significant evidence of heterogeneity. Statistical analysis was performed using the Review Manager software version 5.4. The risk of publication bias was assessed using the ROBINS-I^15^ tool for non-randomized studies. The systematic review and meta-analysis were carried out in accordance with the Cochrane^16^ guidelines and the PRISMA^17^ 2020 declaration for the preparation of intervention systematic reviews. Data analysis was performed using the Review Manager version 5.4 software to obtain results. Egger’s visual inspection test was not performed due to the inclusion of less than 10 studies^18,19^. The literature review in databases, the variables obtained, the data processing and the results were managed in accordance with the regulations for the protection of sensitive information and confidentiality, and were used exclusively for academic and research purposes.

## RESULTS

In the systematic search of the six databases, duplicate articles were identified and excluded, the inclusion and exclusion criteria were applied by title and/or abstract of the article, and later by review of the full text and during the extraction of the articles data (Figure 1). A total of 558 published articles were found, of which only three met the inclusion criteria and were submitted to analysis, with a total sample of 69 patients^20–22^. The average age was 62 years, being predominantly female (79%). Table 1 shows the baseline demographic characteristics of the population in each study. One third of the population had a history of dyslipidemia and diabetes. The largest population with coronary artery disease was found in the study by Vitsenya et al (33 %), where 42.9 % also had some known arrhythmia (predominantly atrial fibrillation). The 40 % (Vitsenya et al) and 33.3 % (Frey et al) of the patients had arterial hypertension at the time of inclusion in the studies. Table 2 shows the concomitant therapy for the management of heart failure in the population of each study. The entire population received some ACEi or ARB in the three studies; in the studies by Vitsenya et al and Frey et al, they are reported as 100 % for both pharmacological groups, however, it is specified that it was one or the other drug, never both at the same time. Between 85 % and 100 % of the patients received some BB. More than half of the patients received an MRA. The use of diuretics was present from 47 % to 71 % of the patients. Less than a third of the populations in the studies by Gregorietti et al and Frey et al had an implantable cardioverter-defibrillator (ICD) or Cardiac Resynchronization Therapy-Defibrillation (CRT-D).

**Table 1.** Baseline Characteristics of the Study Population. CAD: Coronary Artery Disease; SAH: Systemic Arterial Hypertension; AF: Atrial Fibrillation. (*) or another arrhythmia. The boxes with (-) indicate that this variable was not reported in the corresponding study.

| Baseline Characteristics of the Study Population |  |  |  |  |  |  |  |  |
| --- | --- | --- | --- | --- | --- | --- | --- | --- |
| Trial | Age (years) | Female n (%) | Male n (%) | Dyslipidemia n (%) | Diabetes n (%) | CAD n (%) | SAH n (%) | AF* n (%) |
| Gregoriotti 2020 | 56.2 +/- 13.4 | 25 (89.3) | 3 (10.7) | 10 (35.71) | 8 (28.6) | 2 (7.14) | - | - |
| Vitsenya 2020 | 61 (51-67) | 20 (100) | 0 (0) | - | 1 (5) | 0 (0) | 8 (40) | - |
| Frey 2021 | 70 (20-91) | 10 (48) | 11 (52) | 5 (23.8) | 5 (23.8) | 7 (33.3) | 7 (33.3) | 9 (42.9) |

**Table 2.**
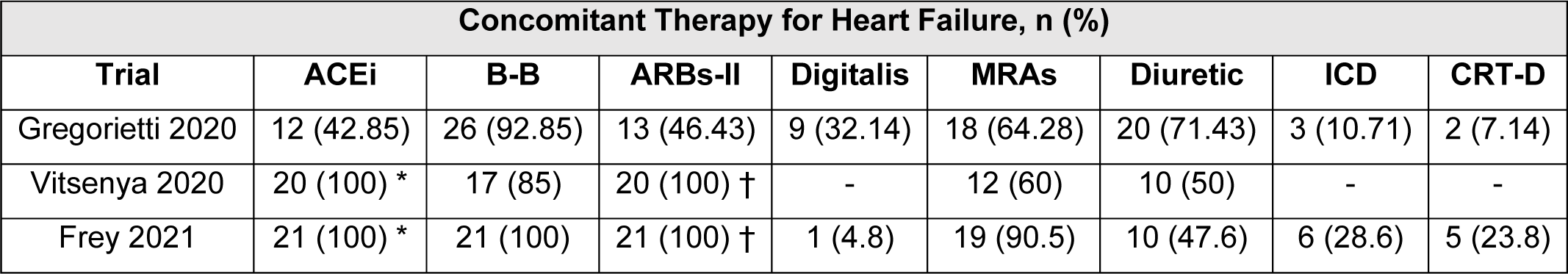
Concomitant Therapy for Heart Failure. ACEi: Angiotensin Converting Enzyme Inhibitor; B-B: Beta Blocker; ARBs-II: Angiotensin Receptor Blocker type II; MRAs: Mineralocorticoid Receptor Antagonist; ICD: implantable cardioverter-defibrillator; CRT-D: Cardiac Resynchronization Therapy-Defibrillation. (\**)= or ARBs-II, (*†*)= or ACEi*. The boxes with (-) indicate that this variable was not reported in the corresponding study.

| Concomitant Therapy for Heart Failure, n (%) |  |  |  |  |  |  |  |  |
| --- | --- | --- | --- | --- | --- | --- | --- | --- |
| Trial | ACEi | B-B | ARBs-II | Digitalis | MRAs | Diuretic | ICD | CRT-D |
| Gregoriotti 2020 | 12 (42.85) | 26 (92.85) | 13 (46.43) | 9 (32.14) | 18 (64.28) | 20 (71.43) | 3 (10.71) | 2 (7.14) |
| Vitsenya 2020 | 20 (100) * | 17 (85) | 20 (100) † | - | 12 (60) | 10 (50) | - | - |
| Frey 2021 | 21 (100) * | 21 (100) | 21 (100) † | 1 (4.8) | 19 (90.5) | 10 (47.6) | 6 (28.6) | 5 (23.8) |

The effects of the use of sacubitril/valsartan on variables that assess ventricular remodeling (echocardiographic and biochemical), functional capacity, and safety profile were analyzed.

## 1. EFFECTS ON VENTRICULAR REMODELING

### 1.1 Percent Change in Left Ventricular Ejection Fraction (LVEF)

All 3 studies examined the effects of sacubitril/valsartan use on the percentage change in left ventricular ejection fraction. A fixed effects model was used due to heterogeneity between studies (I^2^= 67 %). There was a statistically significant percentage increase in LVEF with the use of sacubitril/valsartan [mean difference 6.78 % (95 % CI: 4.91 – 8.65), p < 0.001] compared to baseline LVEF (Figure 1.1).

### 1.2 Percent Change in Left Ventricular Global Longitudinal Strain (LV-GLS)

Two studies examined the effects of the use of sacubitril/valsartan on the change in the percentage of global longitudinal strain of the left ventricle. A fixed effects model was used due to heterogeneity between studies (I^2^= 39 %). There was a statistically significant percentage increase in LV-GLS with the use of sacubitril/valsartan [mean difference 3.37 % (95 % CI: 1.91 – 4.82), p < 0.001] compared to baseline LV-GLS (Figure 1.2).

### 1.3 Change in left atrial volume (LAV)

Two studies examined the effects of sacubitril/valsartan use on change in left atrial volume. A fixed effects model was used due to the null heterogeneity between studies (I^2^= 0 %). There was a statistically significant decrease in left atrial volume with the use of sacubitril/valsartan [mean difference −9.91 % (95 % CI −14.32 to −5.50), p < 0.001] compared to baseline LAV (Figure 1.3).

### 1.4 Change in Left Ventricular End Diastolic Volume (LVED)

Two studies examined the effects of sacubitril/valsartan use on change in left ventricular end-diastolic volume. A fixed effects model was used due to heterogeneity between studies (I^2^= 54

%). There was a statistically significant decrease in left ventricular end-diastolic volume with the use of sacubitril/valsartan [mean difference −5.67 % (95 % CI −11.02 to −0.32), p = 0.04] compared to baseline LVED (Figure 1.4).

### 1.5 Change in serum NT-proBNP levels

All three studies examined the effects of sacubitril/valsartan use on the change in serum NT-proBNP levels. A fixed effects model was used due to heterogeneity between studies (I^2^= 64 %). There was a statistically significant decrease in serum NT-proBNP levels with the use of sacubitril/valsartan [mean difference of −1003.64 pg/mL (95 % CI: −1334.49 to −672.78), p < 0.001] compared with NT-basal proBNP (Figure 1.5).

## 2. EFFECTS ON FUNCTIONAL CAPACITY

### 2.1 Change in meters walked in the 6-minute walk test (SMWT)

Two studies examined the effects of sacubitril/valsartan use on change in meters walked in the 6-minute walk test. A fixed effects model was used due to heterogeneity between studies (I^2^= 70 %). There was a statistically significant increase in distance walked with the use of sacubitril/valsartan [mean difference 95.23 meters (95 % CI: −1334.49 to −672.78), p < 0.001] compared to baseline SMWT (Figure 2.1).

## 3. EFFECTS ON DRUG SAFETY

### 3.1 Change in serum creatinine levels (mg/dL)

All three studies examined the effects of sacubitril/valsartan use on the change in serum creatinine levels. A fixed effects model was used due to the null heterogeneity between studies (I^2^= 0 %). There were no statistically significant changes with the use of sacubitril/valsartan [mean difference 0.04 mg/dL (95 % CI: −0.02 – 0.11), p = 0.19] compared to baseline creatinine levels (supplementary Figure 3.1).

### 3.2 Change in serum potassium levels (mmol/L)

All three studies examined the effects of sacubitril/valsartan use on changing serum potassium levels. A fixed effects model was used due to the null heterogeneity between studies (I^2^= 0 %). There were no statistically significant changes with the use of sacubitril/valsartan [mean difference 0.08 mg/dL (95 % CI: −0.00 – 0.16), p = 0.05] compared to baseline potassium levels (supplementary Figure 3.2).

## DISCUSSION

Cardiovascular disease has become one of the leading causes of mortality in long-term cancer survivors. In this group of patients, early-onset cardiotoxicity is a strong predictor of late-onset or chronic cardiomyopathy. Heart failure due to cardiomyopathy induced by anthracyclines is a fatal complication, reaching up to 60 % mortality rate at two years. Regarding the use of sacubitril/valsartan in patients with heart failure secondary to the use of anthracyclines, to date there are only non-randomized prospective cohorts and series of case reports, however, there are no randomized clinical trials evaluating its beneficial effect in this population. Therefore, to our knowledge, this is the first meta-analysis evaluating the response to treatment with sacubitril/valsartan in patients with anthracycline cardiotoxicity. In the population analyzed in our study, the presence of comorbidities such as dyslipidemia, diabetes, coronary artery disease, and arterial hypertension stand out, as they are known risk factors for developing anthracycline-induced cardiomyopathy^23^. In our study, the female sex predominated because breast cancer was the most prevalent in the three studies, and it is the main cancer in frequency, followed by hematological cancer, with use of anthracyclines. Angiotensin-II receptor plays an important role in anthracycline-induced cardiomyopathy, by increasing oxidative stress that leads to structural damage to cardiomyocytes^24^. We can see that in the population analyzed, all the patients were managed with some ACEi or ARB. The only pharmacological group that is not mentioned in the studies are the sodium-glucose cotransporter type 2 inhibitors, perhaps due to the little existing evidence at the time of their performance, but which we now know is part of the treatment pillars. In evaluating myocardial function, LVEF has been shown to be an independent predictor of short- and long-term mortality in patients with anthracycline-induced cardiomyopathy,^25^ so that its decrease (symptomatic or asymptomatic) is associated with higher mortality in these patients^26^. For this reason, LVEF was the main variable in the evaluation of myocardial function in our study, which included those patients with a subclinical reduction in systolic function. When analyzing the effects of the use of sacubitril/valsartan on LVEF, there was an increase with respect to the baseline, which translates into a quantitative improvement with clinical implication since there was improvement in the functional class; however, due to the way this last variable was reported, it was not possible to perform the analysis of paired groups. A meta-analysis analyzed different potentially cardioprotective therapies, of which only dexrazoxane^27^ was shown to reduce the probability of heart failure; however, it did not show a benefit on survival. We must take into account that the United States Food and Drug Administration (FDA) and the European Medical Agency limit the approval of the use of dexrazoxane for patients treated with high-dose anthracyclines, who are with metastatic disease and not in the adjuvant setting. With the use of beta-blockers and angiotensin blockers^28,29^, there was an improvement of 6.06 %; however, no difference was observed between the primary and secondary outcomes according to the number of cardioprotective agents (monotherapy versus combination), so in our study, the concomitant use of other drugs might not contribute to the effect, however this finding is just suggestive at best and in need of formal investigation. Other authors^30,31^ reported that the use of ACEi/ARB preserved the LVEF compared with controls, but Gujral et al^32^ found no significant difference.

Regarding the use of beta-blockers^33,34^, there is a benefit for drop in LVEF, with a significant increase in LVEF reported in favor of beta-blockers at maximum 4.46 %. However, Huang et al^35^ reported a smaller improvement in LVEF (3.47 %) and the sensitivity analysis excluding the study with the greatest heterogeneity (the study that did not report the standard deviation of LVEF) lost statistical significance. About statins, there are reported an maximum improvement in LVEF of 6.14 % with the use of statins^36,37^. For its part, sacubitril/valsartan in this meta-analysis demonstrated an improvement in LVEF of 6.78 %, being one of the highest values documented to date.

Since BNP responds to end-diastolic pressure and NT-proBNP is more sensitive, this variable was analyzed in our study. Slight increases in serum BNP levels identify a risk of cardiac dysfunction, suggesting that elevated BNP levels may be a diagnostic marker of cardiotoxicity risk and even improve diagnostic performance of another similar cardiac dysfunction related to cancer therapy. In our study, a reduction in serum levels was found after the use of sacubitril/valsartan, which suggests an improvement in diastolic function, which could be a useful index for monitoring in the early stages of cardiotoxicity. There is not improvement for the BNP variable with the use of carvedilol combined with candesartan^34^.

The strength of our study is that analyzes of variables other than LVEF were included, since it has been documented that a drop in LVEF is not the only finding indicative of cardiotoxicity.

Another question is whether there is a difference in the rate of response to cardioprotective drugs according to gender, so we did a sensitivity analysis finding an improvement in LVEF in women of 7.46 %, so it will be necessary to investigate whether sacubitril/valsartan has a greater benefit in women or is the higher prevalence of breast cancer the confounding factor. The inclusion of observational studies potentially strengthened our conclusions by allowing longer follow-up.

The main finding of this analysis of three non-randomized prospective studies is that in patients with heart failure attributable to the use of anthracyclines, the use of sacubitril/valsartan preserves and even increases the percentage of LVEF after chemotherapy-induced damage.

### Study limitations

Possible confounding factors such as race and smoking status were not included in the report, because we did not have access to these data, so it was not possible to adjust the results in this meta-analysis. Dose of anthracycline used was unknown, so it was not possible to perform a subgroup analysis for the different doses. Due to the small number of studies, we were unable to assess publication bias or perform meta-regression or another exploration of heterogeneity.

## CONCLUSIONS

This meta-analysis suggests that there is benefit on the variables that evaluate ventricular remodeling and functional capacity, with the use of sacubitril/valsartan in patients with heart failure related to the use of anthracyclines, without altering the safety profile measured by serum creatinine and potassium levels. This is the first meta-analysis to our knowledge evaluating the effect of sacubitril/valsartan in patients with anthracycline-induced cardiotoxicity. Randomized double-blind placebo-controlled clinical trials are required to confirm these findings.

## Data Availability

There are no extraordinary data to be shown other than those mentioned in the manuscript.

## ACKNOWLEDGMENTS

The authors declare that there are not acknowledgments in this paper.

---

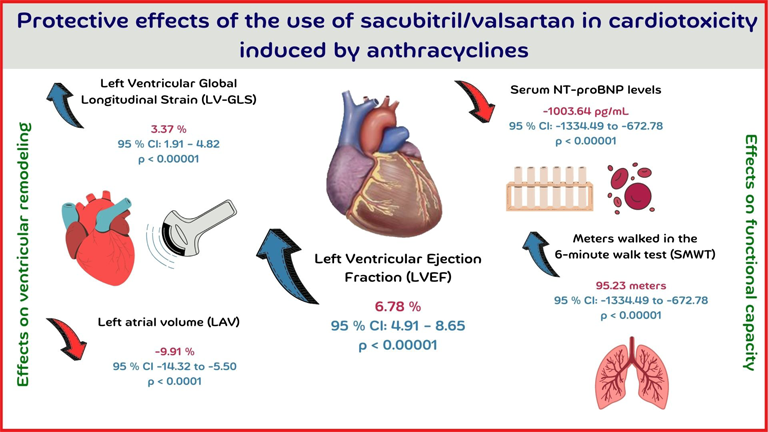

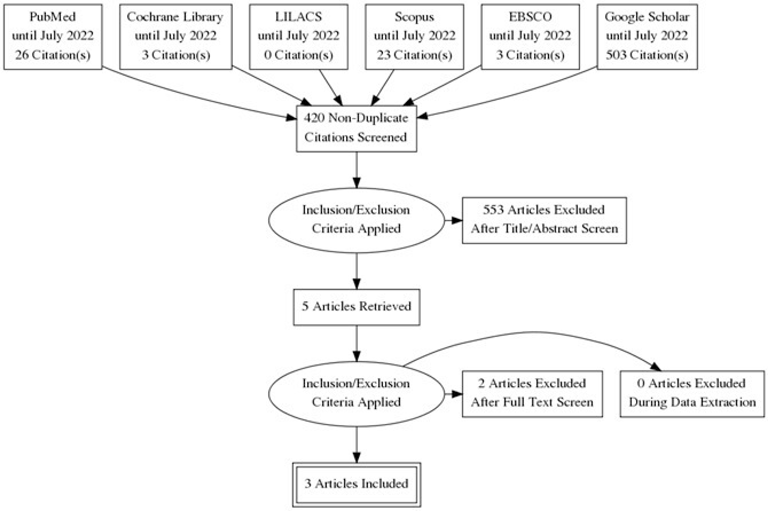

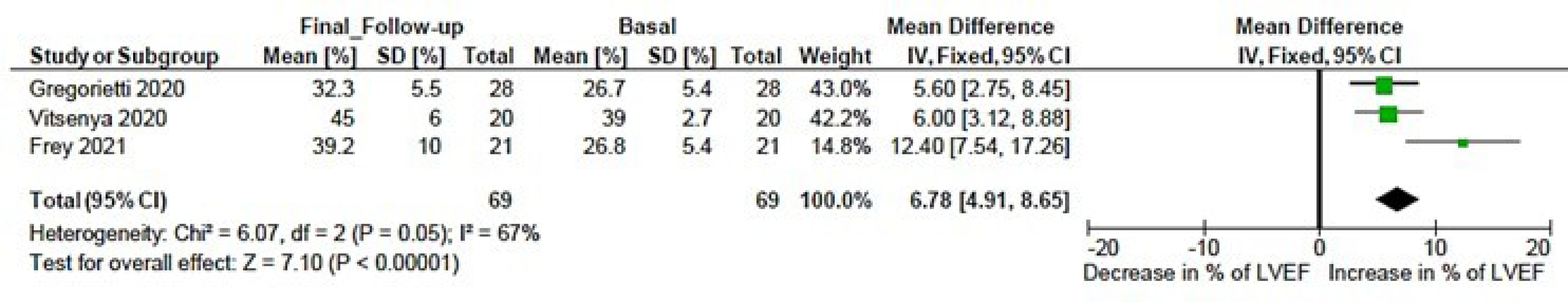

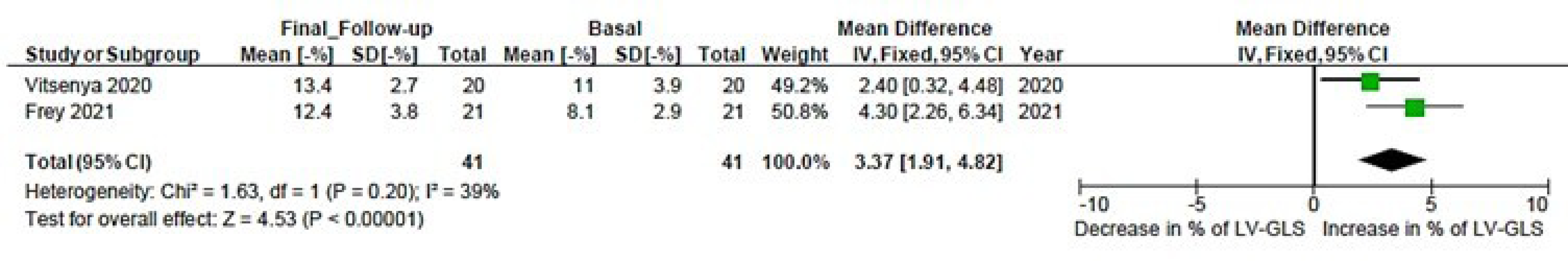

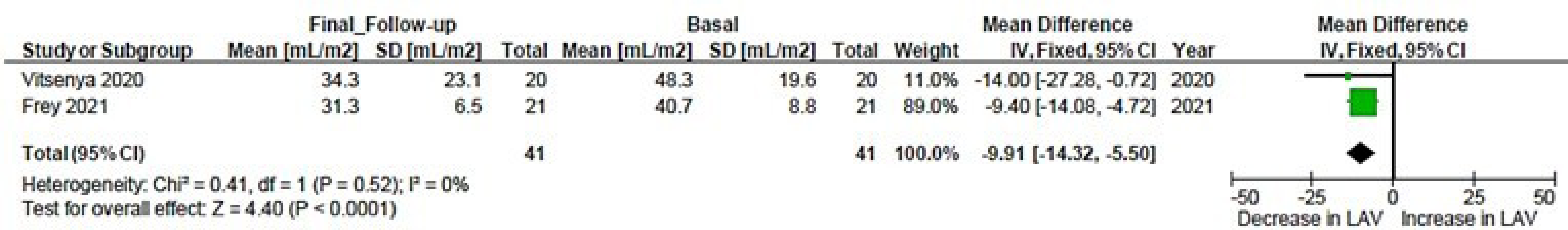

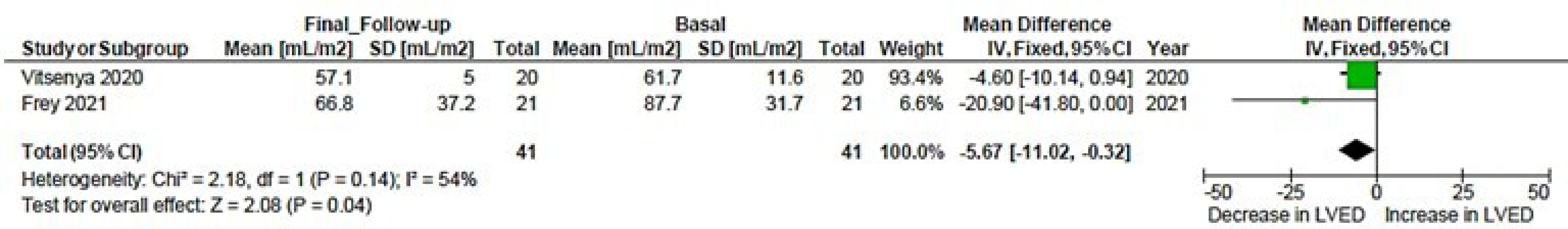

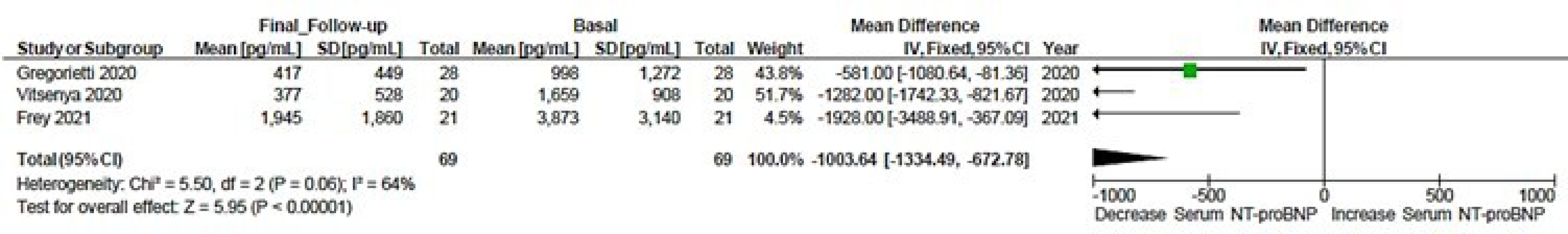

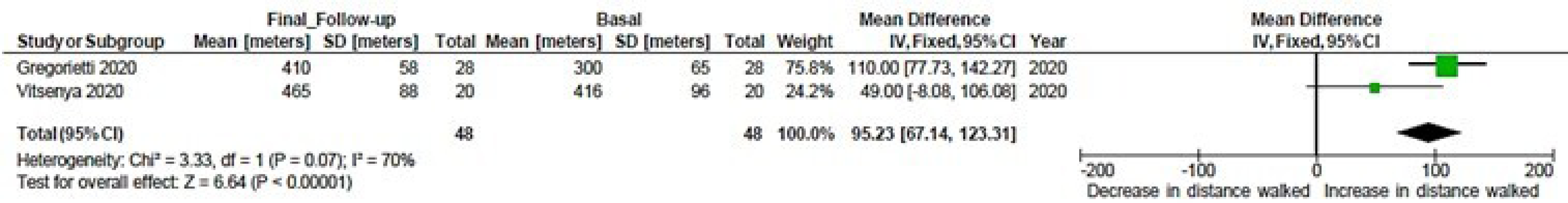

